# Detection of bacterial co-infections and prediction of fatal outcomes in COVID-19 patients presenting to the emergency department using a 29 mRNA host response classifier

**DOI:** 10.1101/2022.03.14.22272394

**Authors:** Nikhil Ram-Mohan, Angela J. Rogers, Catherine A. Blish, Kari C. Nadeau, Elizabeth J Zudock, David Kim, James V. Quinn, Lixian Sun, Oliver Liesenfeld, Stanford COVID-19 Biobank Study Group, Samuel Yang

## Abstract

**Objective:** Clinicians in the emergency department (ED) face challenges in concurrently assessing patients with suspected COVID-19 infection, detecting bacterial co-infection, and determining illness severity since current practices require separate workflows. Here we explore the accuracy of the IMX-BVN-3/IMX-SEV-3 29 mRNA host response classifiers in simultaneously detecting SARS-CoV-2 infection, bacterial co-infections, and predicting clinical severity of COVID-19.

**Methods:** 161 patients with PCR-confirmed COVID-19 (52.2% female, median age 50.0 years, 51% hospitalized, 5.6% deaths) were enrolled at the Stanford Hospital ED. RNA was extracted (2.5 mL whole blood in PAXgene Blood RNA) and 29 host mRNAs in response to the infection were quantified using Nanostring nCounter.

**Results:** The IMX-BVN-3 classifier identified SARS-CoV-2 infection in 151 patients with a sensitivity of 93.8%. Six of 10 patients undetected by the classifier had positive COVID tests more than 9 days prior to enrolment and the remaining oscillated between positive and negative results in subsequent tests. The classifier also predicted that 6 (3.7%) patients had a bacterial co-infection. Clinical adjudication confirmed that 5/6 (83.3%) of the patients had bacterial infections, i.e. *Clostridioides difficile* colitis (n=1), urinary tract infection (n=1), and clinically diagnosed bacterial infections (n=3) for a specificity of 99.4%. 2/101 (2.8%) patients in the IMX-SEV-3 Low and 7/60 (11.7%) in the Moderate severity classifications died within thirty days of enrollment.

**Conclusions:** IMX-BVN-3/IMX-SEV-3 classifiers accurately identified patients with COVID-19, bacterial co-infections, and predicted patients’ risk of death. A point-of-care version of these classifiers, under development, could improve ED patient management including more accurate treatment decisions and optimized resource utilization.

## Introduction

Clinicians in the Emergency Department (ED) face major challenges in accurately assessing patients with suspected infections including SARS-CoV-2, bacterial co-infections, as well as predicting clinical outcomes. Bacterial co-infections (at presentation) and superinfections (after presentation)^1,2^ often cause worse outcomes than the primary viral infection^3^; this phenomenon was prevalent in the H1N1 influenza pandemic^4^, with 20% -30% bacterial coinfections in patients with severe influenza^5,6^. However, current evidence for COVID-19 portrays a different scenario. Recent studies have shown rates of bacterial co-infection in COVID-19 of between 3.2% and 5.5%^1,7–9^, with rates of secondary or superinfection in hospitalized patients increasing to 6.1% -15%^1,7,10,11^. Despite the relatively low prevalence of bacterial co-infections in COVID-19, empiric antibiotics for community or hospital acquired bacterial pneumonia or bacteremia are often prescribed in severely ill patients due to the inability to accurately or rapidly detect bacterial co-infection at presentation^1,12,13^.

Existing diagnostic tests have major limitations. Gold standard bacterial cultures often take days to result, are limited by the ability of the organism to grow in the culture medium, and require a large sample volume when testing complex patient samples like blood^14,15^. Additionally, false negatives can result from insufficient culture duration, or antimicrobial treatment prior to sample collection^16^. False negative culture results can have devastating consequences for patients. Alternate testing methods involve polymerase chain reaction based (PCR) targeted amplification of bacterial nucleic acids directly from the patient’s blood sample. These are not routinely used in the acute setting, are limited by turnaround time and the panel of targets they can detect, and are influenced by the inherent issues of PCR – lack of sensitivity in detecting low bacterial loads, sensitivity to protocols and threshold decisions adopted, and the presence of inhibitory molecules in complex samples such as blood^17^.

There is therefore an unmet medical need to identify viral and bacterial infection using rapid point-of-care tests in the ED to determine presence and severity of infection and inform the use of antimicrobials. In the absence of such diagnostics, clinical decision making needs to balance antimicrobial stewardship with delivery of appropriate empiric care, including escalation of therapy in patients with suspected bacterial co-infections and/or suspected sepsis, and to predict severity for level of care decisions, and optimal use of healthcare resources.

The machine-learning supported host response mRNA classifier IMX-BVN-2 has recently been described to accurately identify systemic as well as localized bacterial infections and also viral infections other than COVID-19^18^. A separate classifier, IMX-SEV-2, has been developed to predict the illness severity (Galtung et al, in revision). The identity and biological functions of the 29 host mRNAs have recently been published^19^, and the classifiers have been further updated (IMX-BVN-3 and IMX-SEV-3) based on additional clinical study data.

The aim of this study was to investigate the accuracy of IMX-BVN-3 and IMX-SEV-3 classifiers to detect SARS-CoV-2 infection, detect bacterial co-infections and predict the severity in patients with confirmed COVID-19.

## Methods

### Patient enrollment and specimen collection

One hundred sixty-one patients with PCR-confirmed COVID-19 infection at presentation were enrolled at the Emergency Department of Stanford University Hospital, USA under the IRB approved protocols 55650 and 55924. 2.5 mL of whole blood was collected in PAXgene Blood RNA tubes (PreAnalytiX) within 12 hours of presenting to the ED and frozen following the instructions of the manufacturer.

Clinical data collected, in the form of a structured questionnaire, included presence of symptoms, past medical history, medications, hospital length of stay (hours and days), CRP, procalcitonin, LDH, and ferritin levels, and neutrophil, lymphocyte, monocyte, eosinophil, and basophil counts. In addition, we determined the patient’s clinical outcomes in the form of disposition from the Emergency Department, need for mechanical ventilation, and death.

### PAXgene sample processing

PAXgene Blood RNA (PreAnalytiX, Hombrechtikon, Switzerland) tubes were shipped to Inflammatix Inc. (Burlingame, CA) under a sponsored research agreement where RNA was extracted using a protocol previously described^20^ and 29 host mRNAs were quantified using the nCounter FLEX instrument (Nanostring, Seattle, WA).

### IMX-BVN-3 and IMX-SEV-3 classifiers

Quantification results for the 29 host mRNAs were analyzed using the BVN-3 and SEV-3 host response classifiers. The classifiers generate numerical scores for the likelihood of bacterial infection and the likelihood of viral infection that each fall into 4 diagnostic (Very unlikely, Unlikely, Possible, Very likely bacterial and/or viral infection) and a score for the condition’s severity that falls into three prognostic interpretation bands (Low, Moderate, and High severity).

### SARS-CoV-2 RNA quantification

Plasma and nasopharyngeal viral RNA levels in Cycle threshold (Ct) and absolute copies/uL were determined for 89/161 COVID-19 positive patients co-enrolled in our previous study^21^ to correlate viral load with the likelihood scores. Briefly, RNA was extracted from 140 μL of samples using the QIAamp Viral RNA Mini Kit (Qiagen, Germany) and quantified using the |Q| Triplex Assay with the qPCR platform QuantStudio 5 (Applied Biosystems by Thermo Fisher Scientific) and dPCR using the array-based |Q| assay simultaneously.

### Clinical adjudication of bacterial co-infections

Blood for culturing was collected from 58/161 patients suspected of an infection. Blood culture results and labs were compared against the IMX-BVN-3 bacterial likelihood scores. A thorough chart review was performed on patients with discordant IMX-BVN-3 bacterial likelihood scores and bacterial culture results and other laboratory results to identify any patient with suspected bacterial infection. Bacterial infection was confirmed if the patient had: 1) ED/inpatient primary or relevant discharge diagnoses that included sepsis, septic shock, or any bacterial infections, 2) positive microbiological data for bacterial pathogens collected within 48 hours from ED presentation, or 3) infectious disease expert consultation documenting bacterial infection upon hospital admission.

### Statistical analysis

We calculated the Pearson correlation between the IMX-BVN-3/IMX-SEV-3 viral likelihood scores and severity with the absolute viral load in the nasopharynx and plasma for 89 patients described elsewhere^21^, between the bacterial likelihood scores and levels of C-reactive protein, procalcitonin, and lactate dehydrogenase, and the Spearman rank correlation between the Cycle threshold (Ct) and the viral likelihood scores. We compared the viral loads between the true positive and false negative calls of viral infection as well as the severity scores between clinical outcomes using the Wilcoxon rank sum test with continuity correction and adjusted the p-value when comparing multiple outcomes using the Benjamin and Hochberg correction. We also calculated the sensitivity, specificity, and the likelihood ratios of the viral and bacterial classification bands against the PCR COVID-19 positivity and adjudicated bacterial co-infections respectively. Additionally, we also compared the proportions of patients in the severity likelihood bands and their clinical outcomes – disposition from the ED and the need for ventilation/30-day mortality using χL2 tests with continuity corrections. All analyses were performed in R.

## Results

### Patient characteristics

A total of 161 patients were enrolled from April 2020 to February 2021, with median age of 50 years (IQR: 35 -64). 84/161 (52.2%) were women. 158/161 (98.1%) were symptomatic on presentation with a median of 6 symptoms (IQR: 4 -8). Medical history, comorbidities and symptoms at presentation are shown in **Table 1**.

**Table 1.**
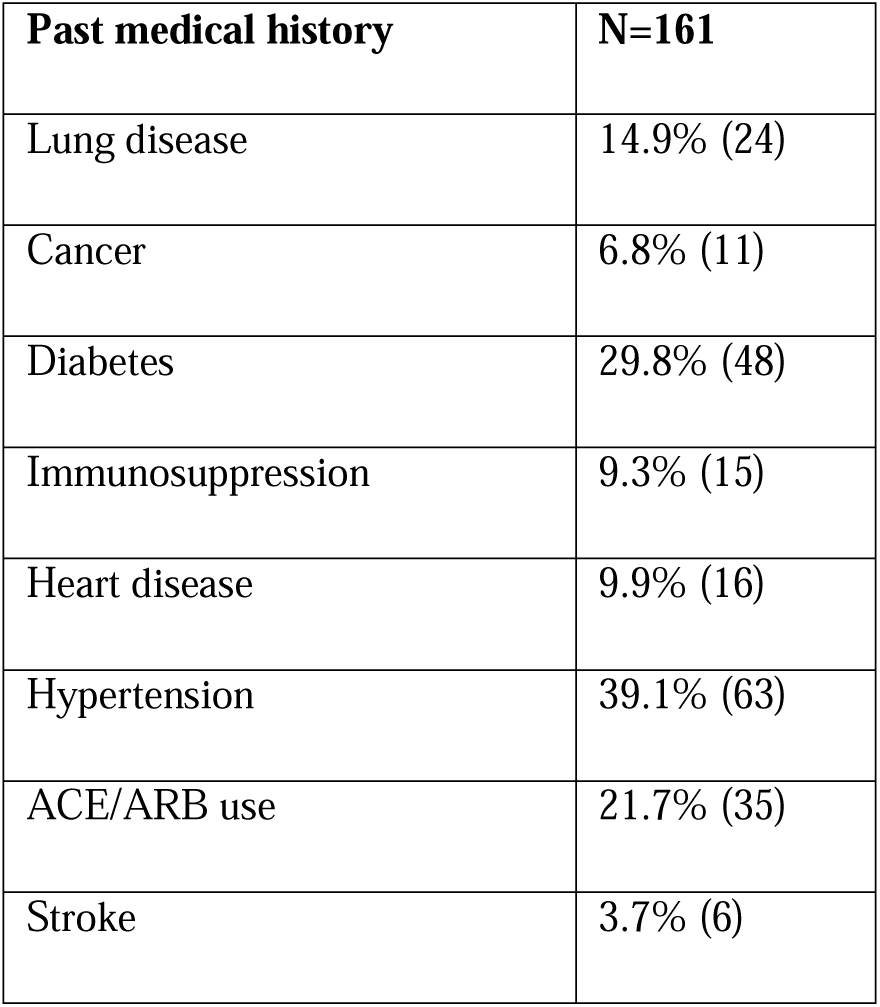
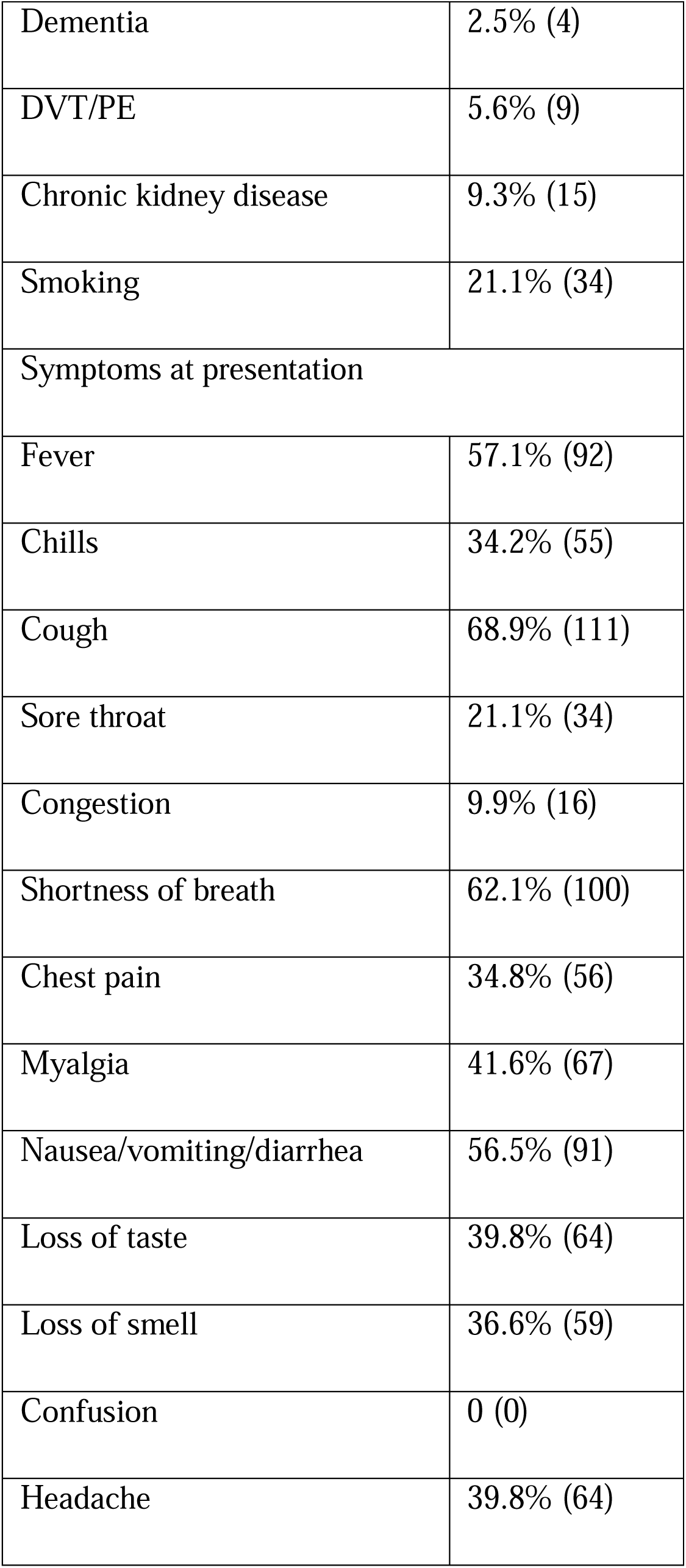
Patient medical history and symptoms at presentation

| Past medical history | N=161 |
| --- | --- |
| Lung disease | 14.9% (24) |
| Cancer | 6.8% (11) |
| Diabetes | 29.8% (48) |
| Immunosuppression | 9.3% (15) |
| Heart disease | 9.9% (16) |
| Hypertension | 39.1% (63) |
| ACE/ARB use | 21.7% (35) |
| Stroke | 3.7% (6) |
| Dementia | 2.5% (4) |
| DVT/PE | 5.6% (9) |
| Chronic kidney disease | 9.3% (15) |
| Smoking | 21.1% (34) |
| Symptoms at presentation |  |
| Fever | 57.1% (92) |
| Chills | 34.2% (55) |
| Cough | 68.9% (111) |
| Sore throat | 21.1% (34) |
| Congestion | 9.9% (16) |
| Shortness of breath | 62.1% (100) |
| Chest pain | 34.8% (56) |
| Myalgia | 41.6% (67) |
| Nausea/vomiting/diarrhea | 56.5% (91) |
| Loss of taste | 39.8% (64) |
| Loss of smell | 36.6% (59) |
| Confusion | 0 (0) |
| Headache | 39.8% (64) |

### Accuracy in predicting COVID-19 infection using host response markers

151/161 (93.8%) of patients positive for COVID-19 by RT-PCR were accurately classified as “Possible” or “Very Likely” viral infection by IMX-BVN-3, corresponding to an overall sensitivity of 93.8% (85.3% and 7.5% for the Very likely and Possible viral bands, respectively; **Table 2**). As all patients were confirmed SARS-CoV-2 positive we did not calculate specificity of the classifier.

**Table 2.** Break down of patients into viral likelihood interpretation bands using IMX-BVN-3*

| **IMX-BVN-3 viral**<br>**interpretation**<br>**band** | **Frequency** | **Sensitivity**<br>**for COVID-**<br>**19**<br>**(%)** |
| --- | --- | --- |
| Very Likely | 139 | 86.3 |
| Possible | 12 | 7.5 |
| Unlikely | 5 | - |
| Very Unlikely | 5 | - |

\*, specificity was not calculated as all patients were confirmed SARS-CoV-2 positive by PCR

We further investigated the causes of 10 potentially “false negative” results in BVN-3; six of the 10 patients had first tested positive for SARS-CoV-2 at least 9 days before presentation to the ED, while the remaining four had SARS-CoV-2 PCR test results that were initially positive but oscillated between positive and negative when retested. Of interest, 3 of the 10 patients were predicted to have a bacterial superinfection as indicated by the BVN-3 classifier’s bacterial score and 2/3 were clinically adjudicated to have a bacterial infection by expert chart review (see below).

The viral likelihood score was inversely correlated with the PCR cycle threshold value (Ct) from nasopharyngeal samples collected on admission (Spearman rank correlation: -0.63, p<0.001) and correlated with the absolute viral load (copies/uL) as determined by digital PCR (Pearson correlation: 0.52, p< 0.001) (**Figure 1**). Patients with Very Likely or Possible positive BVN-3 viral scores indicating viral infection (“true positives”) had a median viral load of 3,483 copies/μL in the nasopharyngeal sample (IQR: 155 – 23,539) compared to 3.52 copies/μL (IQR: 2.82 – 4.9) in the Unlikely and Very Unlikely BVN-3 (“false negative”) patients (p-value = 0.009).

**Figure 1.**
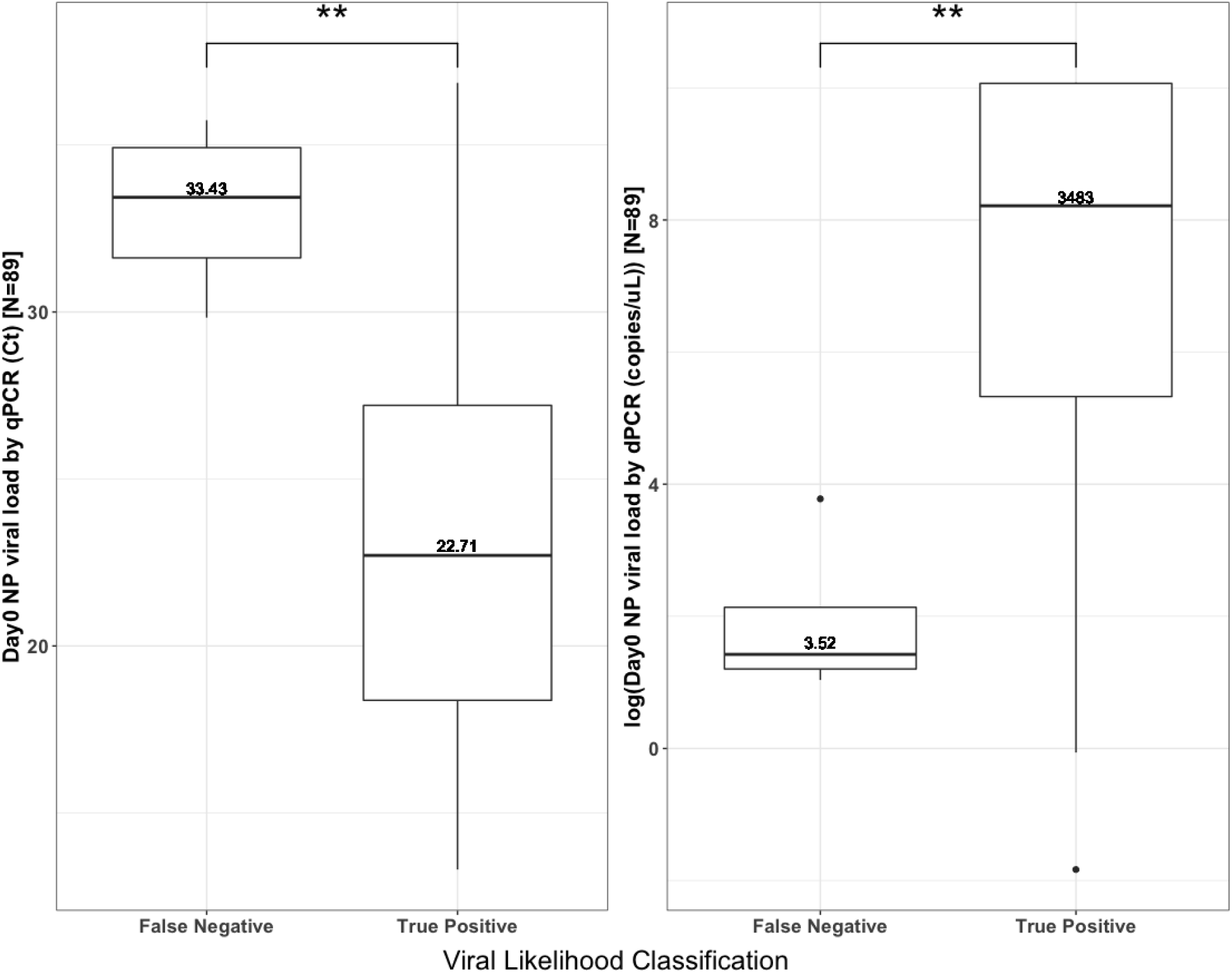
Difference in the nasopharyngeal SARS-CoV-2 load between “false negative” (Unlikely or Very unlikely viral BVN-3 scores) and “true positive” (Possible or Very Likely BVN-3 scores) for 89 patients. qPCR measured viral loads in cycle threshold (Ct) *(left)* and dPCR measured absolute viral loads in copies/μL *(right)*. ** represents p-value < 0.001.

### Detection of bacterial superinfections using host response markers

The IMX-BVN-3 bacterial score classified 6/161 (3.7%) of patients into the Possible bacterial interpretation band suggesting a bacterial co-infection, and 155/161 (96.3%) were classified as Unlikely or Very Unlikely bacterial infections (**Table 3**). Chart review and clinical adjudication confirmed that 5/6 (83.3%) of the Possible bacterial patients did indeed have superinfections translating into a specificity (ruling in) of 99.4% for identification of bacterial infection: one patient had *Clostridioides difficile* colitis, one had rectal adenocarcinoma with gastrointestinal perforation and abdominopelvic abscess and three had clinically diagnosed superinfections without positive microbiology findings (including blood culture) (**Table 4)**. We did not detect evidence for bacterial infections in 52 patients with negative blood culture results translating into a sensitivity of 100% for ruling out bacterial infection in the subgroup of patients where microbiology data were available. The bacterial scores correlated with the levels of C-reactive protein (Pearson correlation: 0.58, p< 0.001), procalcitonin (Pearson correlation: 0.4, p-value = 0.003), and lactate dehydrogenase (Pearson correlation: 0.42, p= 0.003).

**Table 3.** Breakdown of patients into bacterial likelihood interpretation bands using IMX-BVN-3*

| IMX-BVN-3<br>bacterial<br>interpretation<br>band | Confirmed<br>bacterial<br>infection | No<br>bacterial<br>infection | Percent<br>in band | Sensitivity<br>(%) | Specificity<br>(%) | Likelihood<br>ratio |
| --- | --- | --- | --- | --- | --- | --- |
| Very Likely | 0 | 0 | 0 | n.d. | n.d. | n.d. |
| Possible | 5 | 1 | 3.7 | n.d. | 99.4 | 156 |
| Unlikely | 0 | 59 | 36.6 | 100 | n.d | 0 |
| Very Unlikely | 0 | 96 | 59.6 | 100 | n.d. | 0 |

n.d., not determined \*, complete data to calculate sensitivity were only available in 58 patients

**Table 4.**
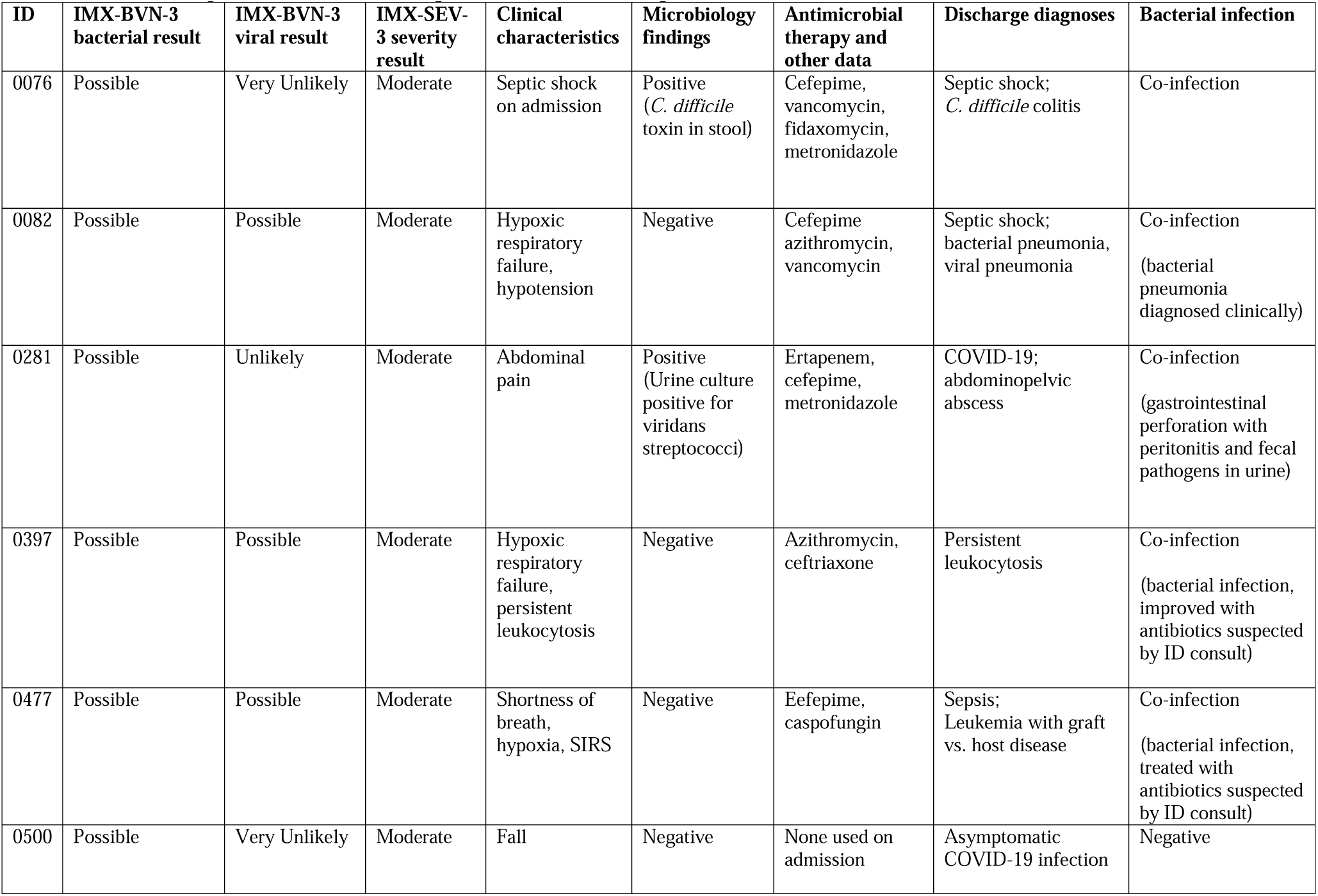
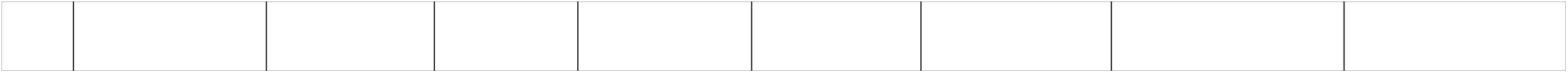
Clinical adjudication based on expert chart review in 6 patients with Possible IMX-BVN-3 bacterial scores

| ID | IMX-BVN-3 bacterial result | IMX-BVN-3 viral result | IMX-SEV-3 severity result | Clinical characteristics | Microbiology findings | Antimicrobial therapy and other data | Discharge diagnoses | Bacterial infection |
| --- | --- | --- | --- | --- | --- | --- | --- | --- |
| 0076 | Possible | Very Unlikely | Moderate | Septic shock on admission | Positive (*C. difficile* toxin in stool) | Cefepime, vancomycin, fidaxomicin, metronidazole | Septic shock; *C. difficile* colitis | Co-infection |
| 0082 | Possible | Possible | Moderate | Hypoxic respiratory failure, hypotension | Negative | Cefepime azithromycin, vancomycin | Septic shock; bacterial pneumonia, viral pneumonia | Co-infection<br>(bacterial pneumonia diagnosed clinically) |
| 0281 | Possible | Unlikely | Moderate | Abdominal pain | Positive (Urine culture positive for viridans streptococci) | Ertapenem, cefepime, metronidazole | COVID-19; abdominopelvic abscess | Co-infection<br>(gastrointestinal perforation with peritonitis and fecal pathogens in urine) |
| 0397 | Possible | Possible | Moderate | Hypoxic respiratory failure, persistent leukocytosis | Negative | Azithromycin, ceftriaxone | Persistent leukocytosis | Co-infection<br>(bacterial infection, improved with antibiotics suspected by ID consult) |
| 0477 | Possible | Possible | Moderate | Shortness of breath, hypoxia, SIRS | Negative | Eefepime, caspofungin | Sepsis; Leukemia with graft vs. host disease | Co-infection<br>(bacterial infection, treated with antibiotics suspected by ID consult) |
| 0500 | Possible | Very Unlikely | Moderate | Fall | Negative | None used on admission | Asymptomatic COVID-19 infection | Negative |

### Disease severity based on host response markers and association with clinical outcomes

The IMX-SEV-3 test classified 101/161 (62.7%) patients in the Low severity category and 60/161 (37.3%) in the Moderate severity category. No patients were categorized in the High severity category. The calculated severity score was correlated with the absolute viral load in plasma (Pearson correlation: 0.49, p-value = 0.002) and the above bacterial score (Pearson correlation: 0.45, p-value < 0.001). Interestingly, 6/6 (100%) patients classified in the Likely bacterial co-infections category were classified to have Moderate severity. The IMX-SEV-3 severity score also correlated with the modified WHO severity score at enrollment for these patients (Pearson correlation: 0.43, p-value < 0.001).

In total, 79/161 (49.1%) patients were discharged, 72/161 (44.7%) patients were admitted to the floor, 10/161 (6.2%) were admitted to ICU, 7/161 (4.3%) required mechanical ventilation (**Table 5**), and 9/161 (5.6%) died. As expected, 59.4% patients in the Low severity category were discharged from the ED compared to only 31.7% in the Moderate category (difference, 27.7% [95% CI: 11.2% -44.2%]) (**Figure 2**). Interestingly, more patients in the Moderate category were admitted to the ICU (difference, 11.4% [95% CI: 1% -21.7%]). Median IMX-SEV-3 severity scores in patients admitted to the ICU were 14.5 (IQR: 13 – 18.25), in those admitted to the floor was 10 (IQR: 8 – 13), and in those discharged it was 8 (IQR: 7 -10). Wilcoxon rank sum test for each pairwise comparison was significant (adjusted p-value < 0.05). When grouping the need for mechanical ventilation and/or mortality as a severe outcome, 13/161 (8.1%) had such a severe outcome from the COVID-19 infection. A greater proportion of patients in the Moderate category had such a severe outcome compared to those in the Low category (15% vs 3.9%, difference, 11.1% [95% CI: 0.09% -22.2%]), and the patients had a higher median IMX-SEV-3 severity score (12 [IQR: 10 -14]) than those that didn’t (9 [IQR: 7 – 11.15], Wilcoxon rank sum test, p-value = 0.07).

**Table 5.**
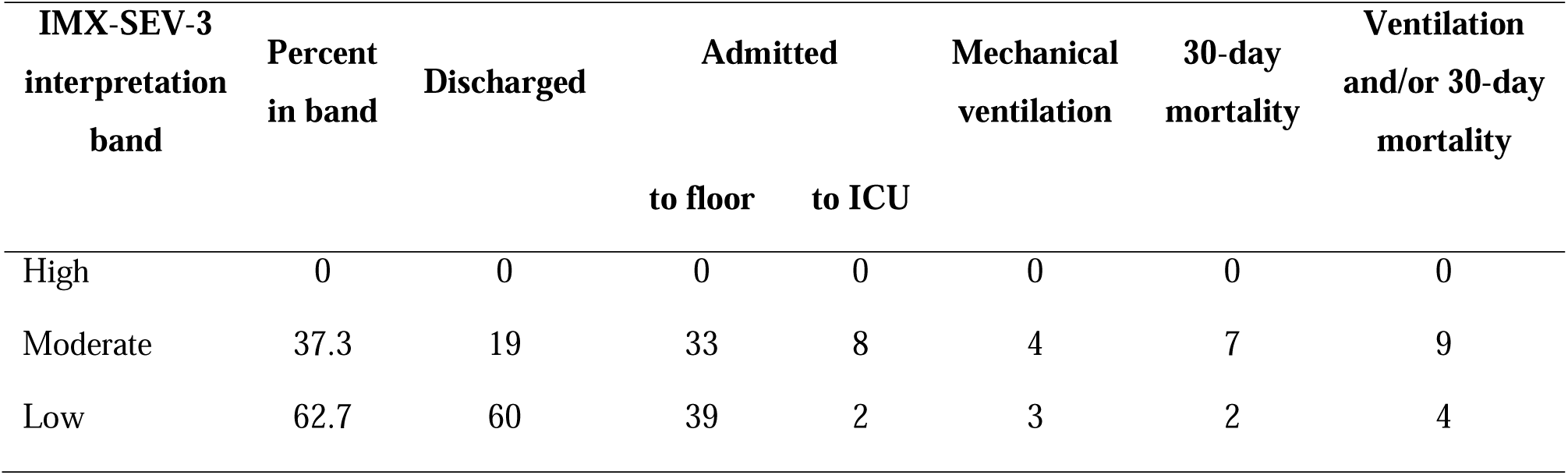
Breakdown of patients into severity interpretation bands using IMX-SEV-3

| IMX-SEV-3<br>interpretation<br>band | Percent<br>in band | Discharged | Admitted |  | Mechanical<br>ventilation | 30-day<br>mortality | Ventilation<br>and/or 30-day<br>mortality |
| --- | --- | --- | --- | --- | --- | --- | --- |
|  |  |  | to floor | to ICU |  |  |  |
| High | 0 | 0 | 0 | 0 | 0 | 0 | 0 |
| Moderate | 37.3 | 19 | 33 | 8 | 4 | 7 | 9 |
| Low | 62.7 | 60 | 39 | 2 | 3 | 2 | 4 |

**Figure 2.**
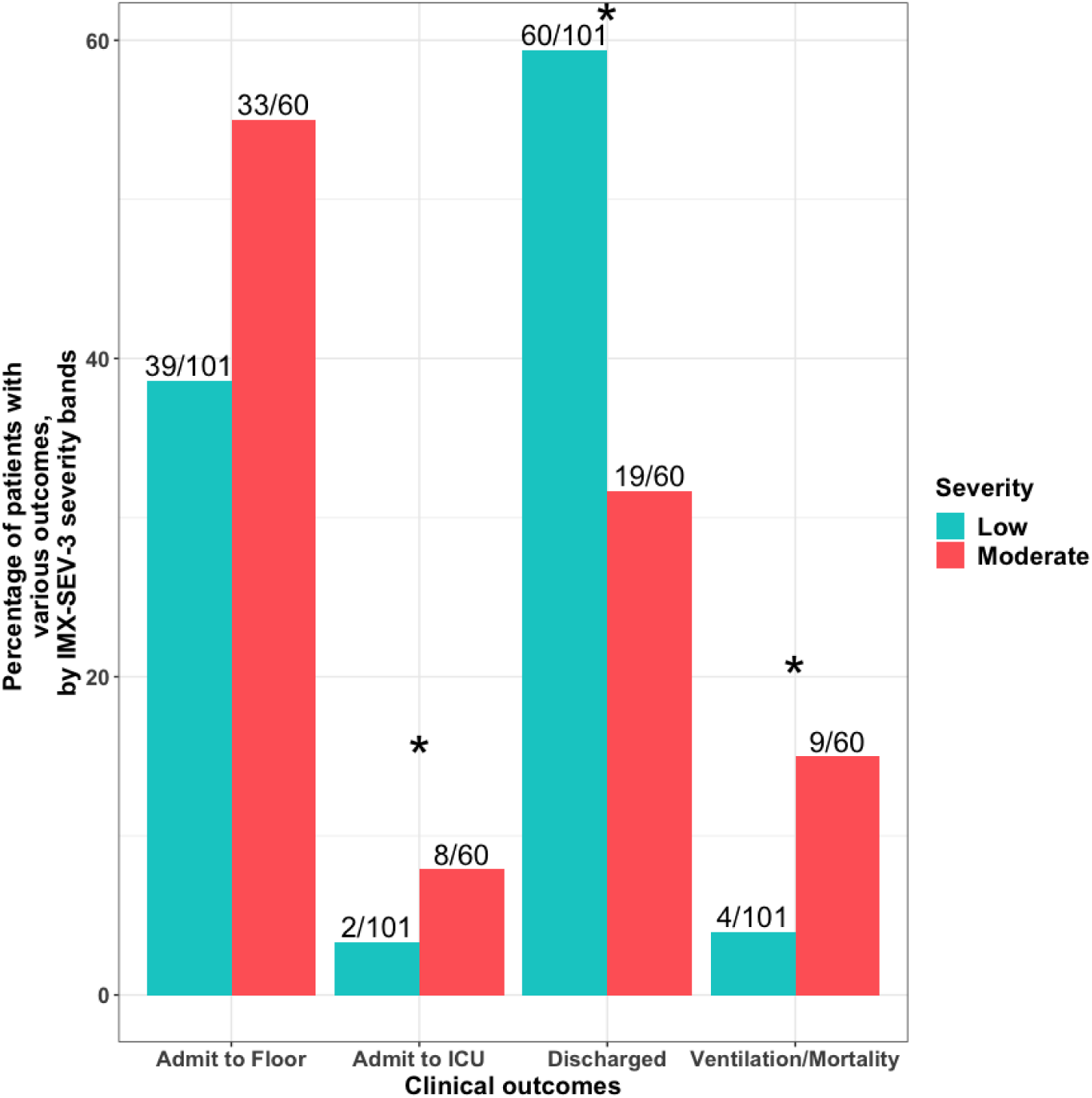
Proportions of patients with different clinical outcomes, including the disposition from the ED as well as the need for ventilation or 30-day mortality, by the severity likelihood predicted by the IMX-SEV-3 classifier. Overall, more patients in the Low category were discharged (difference in proportions, 27.7% [95% CI: 11.2% -44.2%]) and more patients in th Moderate category were admitted to the ICU or required ventilation/succumbed to the infection, difference in proportions of 11.4% [95% CI: 1% -21.7%] and 11.1% [95% CI: 0.09% -22.2%] respectively. * represents p-value < 0.05.

## Discussion

As of January 2022, SARS-CoV-2 has infected more than 340 million people globally and resulted in ∼5.5 million deaths^22^. Bacterial co/superinfections are known to occur in patients infected with SARS-CoV-2 at varying prevalence conditional on the severity of the viral infection^1,7–13,23,24^. Successful detection of the virus requires a high-fidelity PCR test targeting the viral RNA and to date the detection of coinfecting pathogens has depended on either bacterial culture or detecting target nucleic acids in patient samples using PCR. Here we present, to the best of our knowledge, the first host response based simultaneous detection of viral (SARS-CoV-2) infection, co-infection with bacterial pathogens, as well as the stratification of disease severity using the IMX-BVN-3 and IMX-SEV-3 classifiers.

The IMX-BVN-3 classifier detected COVID-19 with 93.8% sensitivity. This is the first report of the successful detection of SARS-CoV-2 using the IMX-BVN-3 host response signature, previously validated in other viral infections^18,25^, and the imputed false negative rate of the classifier is lower than that of the currently accepted qPCR assays for SARS-CoV-2. A recent systematic review and meta-analysis of 32 studies comprised of 18,000 patients revealed heterologous false negative rates in qPCR ranging from 2%^26^ – 58%^27^ with an overall summary estimate of 12%^28^. Of interest, we observed several specific circumstances in the few patients that showed “false negative” results in IMX-BVN-3: first, the time lag between the initial positive SARS-CoV-2 test result and the presentation to the ED in these patients likely indicates clearing of the virus and waning of the associated viral immune response with subsequent negative results in the classifier; second, low viral loads (<5 copies/μL) also contributed to “false-negative” results in the classifier in a few patients. Lastly, two patients with “false negative” classifier results were also found to have bacterial superinfections. As the generation of viral and bacterial scores in the IMX-BVN-3 classifier is interdependent, bacterial scores may have impacted the viral scores and contributed to “false negative” results in addition to the factors mentioned above.

Importantly, the IMX-BVN-3 classifier predicted bacterial co-infection within 48h in 6/161 patients with a specificity of 99.4%. 5/6 were clinically adjudicated to be bacterially infected. We calculated a prevalence of 8.6% (5/58) for bacterial co-infections in a subset of patients with blood cultures available as part of clinical care; this prevalence is similar to the prevalence reported recently for patients with low or moderate SARS-CoV-2 infection^1,7–9^. Importantly, the identification of bacterial co-infections was achieved from the same 2.5 mL blood sample that provided the viral result in IMX-BVN-3 without the need for additional sample collection such as bacterial cultures or samples for the amplification of bacterial nucleic acids. The high accuracy of the IMX-BVN-3 classifier could thus be used along the viral result to initiate antimicrobial treatment and other clinical decisions in SARS-CoV-2 positive patients while also contributing to antimicrobial stewardship in the ED.

The IMX-SEV-3 classifier categorized patients into Low and Moderate severity categories in our cohort. This host-response dependent classifier predicted severity scores that correlated with a modified WHO score that was designed to describe the need for supplemental oxygen^21^. With a significant difference in the median severity scores of patients admitted to the ICU, admitted to the floor, and those who were discharged, as well as the observed increased proportions of patients in the Moderate severity interpretation band admitted to the ICU and having a severe outcome, the severity score could facilitate level of care decision for patients. However, as the study was not powered to assess the accuracy of the severity readout of the IMX-SEV-3 classifier we only report the nominal results here. Additional studies -including a current large registrational trial conducted for clearance by regulatory agencies in the US and Europe-will report the accuracy of the severity readout in larger COVID-19 and other cohorts.

Other limitations of our study include the fact that this study was conducted at a single center and used biobanked blood samples obtained from a limited cohort of 161 patients. As only PCR confirmed COVID-19 positive patients were enrolled we could not determine the IMX-BVN-3 classifier’s specificity. We were also unable to clinically adjudicate the entire patient cohort for bacterial infections and thus calculated sensitivity for a subset of patients only. Lastly, since bacterial co-or superinfections are defined based on when the patient presents to the ED^1,2^ and not when in the course of the infection the patient presents, we were unable to determine the timeline of the infection to distinguish between the two. Additionally, the host response-based classifier detects any bacterial infection and, hence, does not allow differentiating between co-or superinfections.

In conclusion, once the IMX-BVN-3 and SEV-3 classifiers are introduced as a rapid point of care host RNA detection platform with a turnaround time of less than 30 min (currently in development), results at the point of care could guide decisions about starting or withholding antibiotics allowing escalation of therapy or antimicrobial stewardship but also the initiation of contact precaution measures and/or viral therapy and choosing the appropriate level of care for SARS-CoV-2 positive patients.

## Data Availability

All data produced in the present study are available upon reasonable request to the authors

## Acknowledgements

The authors would like to thank the additional author members of the Stanford COVID-19 Biobank Study Group, including Marjan M. Hashemi, Kristel C. Tjandra, Jennifer A. Newberry, Andra L. Blomkalns, Ruth O’Hara, Euan Ashley, Rosen Mann, Anita Visweswaran, Thanmayi Ranganath, Jonasel Roque, Monali Manohar, Hena Naz Din, Komal Kumar, Kathryn Jee, Brigit Noon, Jill Anderson, Bethany Fay, Donald Schreiber, Nancy Zhao, Rosemary Vergara, Julia McKechnie, Aaron Wilk, Lauren de la Parte, Kathleen Whittle Dantzler, Maureen Ty, Nimish Kathale, Arjun Rustagi, Giovanny Martinez-Colon, Geoff Ivison, Ruoxi Pi, Maddie Lee, Rachel Brewer, Taylor Hollis, Andrea Baird, Michele Ugur, Drina Bogusch, Georgie Nahass, Kazim Haider, Kim Quyen Thi Tran, Laura Simpson, Michal Tal, Iris Chang, Evan Do, Andrea Fernandes, Allie Lee, Neera Ahuja, Theo Snow, and James Krempski.

